# Detection of Chikungunya virus in The Gambia through a newly implemented sentinel surveillance program

**DOI:** 10.1101/2024.03.11.24303694

**Authors:** Amadou Woury Jallow, Idrissa Dieng, Bakary Sanneh, Mamadou Aliou Barry, Cheikh Talla, Modou Lamin Sanneh, Samba Niang Sagne, Mamadou Cisse, Alphonse Mendy, Muhammed Kijera, Karamo York, Alieu Faal, Alhagie Papa Sey, Ebrima K. Jallow, Lamin Manneh, Sheriffo M.K. Darboe, Balla Jatta, Momodou Kalisa, Adama M.B. Sanneh, Modou Njie, Momodou T. Nyassi, Mustapha Bittaye, Ndongo Dia, Amadou Alpha Sall, Ousmane Faye, Moussa Moise Diagne, Oumar Faye, Boubacar Diallo, Sheriffo Jagne, Abdourahmane Sow

## Abstract

We characterized 01 autochthonous chikungunya virus (CHIKV) case from (The Gambia) through a newly implemented local arboviruses surveillance program in the, highlighting the first notification of the virus in the Country. Identified virus is closely related to CHIKV West African genotype detected in Kédougou (Senegal) in 2023 and responsible of a large outbreak with up to 300 confirmed cases. This work describes the first genomic proof subregional spread of CHIKV West African genotype in West Africa.

## Background

Emerging infectious diseases, particularly respiratory diseases and arboviruses, pose significant global threats, notably in developing countries (1). The emergence of viral infections has highlighted the need for effective surveillance for early detection of epidemic diseases. Since 2011, both Senegal’s Ministry of Health and Institut Pasteur de Dakar (IPD) have established the Syndromic Sentinel Surveillance Network in Senegal (4S Network), which includes community sites, health centers, and three hospital sites (2,3). The network has reported over 30 outbreaks since 2015 (data not shown), with an increase in arboviruses/viral hemorrhagic fevers detection in 11 regions. This successful model is now planned to be extended to other West African countries through a collaboration between IPD, the West African Health Organization (WAHO) and Africa CDC through the Regional Integrated Surveillance and Laboratory Network RISLNET Project. One of these countries is The Gambia, a neighboring country of Senegal. If the country has already a surveillance system for both malaria (4) and other NTDs (5).

Herein, we describe the setting-up of a 4S-based surveillance system in The Gambia through the RILSNET program and the subsequent detection of the first Chikungunya virus (CHIKV) infection ever reported in the country.

## The study

From January to February 2023, a multidisciplinary team involving virologists and epidemiologists from IPD was deployed in the Gambia to strengthen the surveillance system and the capacity of the staff of the National Public Health Laboratory (NPHL). The main objective was the establishment of two pilots syndromic sentinel surveillance sites in the country for early detection of pathogens and rapid response to outbreaks.

During the process, several meetings were organized with stakeholders of the Ministry of Health (MoH) of the Gambia in order to identify the two sentinel sites following objective selection criteria such as geographic relevance, connectivity with other healthcare facilities and logistic for samples transportation.

During the assessment visit, the heads of each structure were provided with mission terms and procedures for selecting the sentinel sites using a selection tool. On January 23, 2023, at the Bansang Hospital in the upper river region of the Gambia was finally chosen as the first sentinel site in Gambia because of proximity with the Casamance region of Senegal which has experience with arboviruses and respiratory viruses outbreaks. In this site, a training session was conducted on sentinel surveillance procedures, including inclusion criteria, daily epidemiological data transmission, individual case form submission, sample collection and transport, biosecurity and biological safety, and sentinel surveillance equipment was made available. Meanwhile, the National Public Health Laboratory (NPHL) staff benefited a four-day practical training on the reception and diagnosis of suspected cases of arboviruses, respiratory viruses and Hemorrhagic Fever Viruses. At the end of the mission by February 2023, the two syndromic sentinel surveillance sites were set up to enabling them for early detection and investigation of epidemic-potential diseases, with trained staff for real-time data sharing and enhanced collaboration between structures and actors at the central level.

On September 2023, the RISLNET project at two chosen sites conducted a syndromic surveillance system for arboviruses and respiratory infections. The EDC received information from the NPHL confirming a positive case of the chikungunya virus from Bansang General Hospital. The patient was a 11 to 15 -year-old female from the CRR’s and her lab-assigned number was AR/23/85 (which is an anonymized lab number). The EDC informed its partners and stakeholders (NPHL, NMCP, IPD, DHS, DPHS and WHO country Office) of the information when it was received, and it also gave advice to Bansang General Hospital, which treated the patient on August 2023. The EDC assigned the CRR regional surveillance officer to conduct a joint investigation with Bansang General Hospital and produce a report.

### Objectives

According to the investigation, the following objectives were set;

1. To identify the source of the outbreak
2. To determine the magnitude of the outbreak
3. To develop and implement control measures

On late August 2023, the Bansang General Hospital staff detected arbovirus on a girl aged between 11 to 15 years and collected and preserved a blood sample at the hospital’s lab. Along with the blood sample, a case investigation form was completed and delivered to the NPHL. On September 2023, the blood sample was sent to the NPHL via the Sample Referral Network (SRN). On mid September 2023, the sample was analyzed by the NPHL, and it turned positive.

The examined sample was then forwarded to the WHO reference Laboratories on the September 2023 at the Institut Pasteur de Dakar - Senegal (IPD) for confirmatory testing. The sample suggests positive for Chikungunya in this case as well. Four samples, including the positive case, were collected from the positive patient’s close contacts.

## Findings

The case was a girl aged between 11 to 15 years old, case did not travel outside The Gambia. There were no visitors prior to the onset of the disease. Blood Samples were collected and analyzed and the time between sample collection at the Hospital and the turnaround time was found to be three weeks which is not a good practice for infectious disease control. The sample was found to be positive for Chikungunya with CT value of 19.82 using protocol previously described (6) and the sample was sent for confirmation at the reference laboratory at IPD and sample analysis at IPD took less than 12 hours to confirm Chikungunya Positive.

The child had no Infant Welfare Card (IWC) at the time of investigation and during case investigation, four samples were collected from the close contacts and analyzed at the NPHL on September 2023 and all samples including the positive case were negative (Figure S1).

A target enrichment standard hybridization workflow using Twist Biosciences Comprehensive Viral Research Panel (CVRP) was used to obtain the viral whole genome. The RNA extract was reverse transcribed with the SuperScript IV Reverse Transcriptase kit (Invitrogen, Thermo Fisher, USA), and cDNA was fragmented before a telomere repair, a dA-Tailing, a ligation step with Universal Twist adapters before a final libraries amplification as previously described (6). A single pooled library was prepared from the indexed samples, and hybridized targets were bound to streptavidin beads. Enriched sample libraries were loaded onto a Illumina iSeq 100 sequencer, as recommended by the manufacturer.

Phylogenetic analysis showed that the strain identified in Gambia 2023 belongs to the West African genotype of CHIKV and groups with previous sequences from both human and mosquitoes samples collected during an ongoing outbreak in eastern Senegal (Figure 1 ; Phylogenetic tree). highlighting a probable transboundary circulation of CHIKV between Gambia and Senegal.

**Figure 1:**
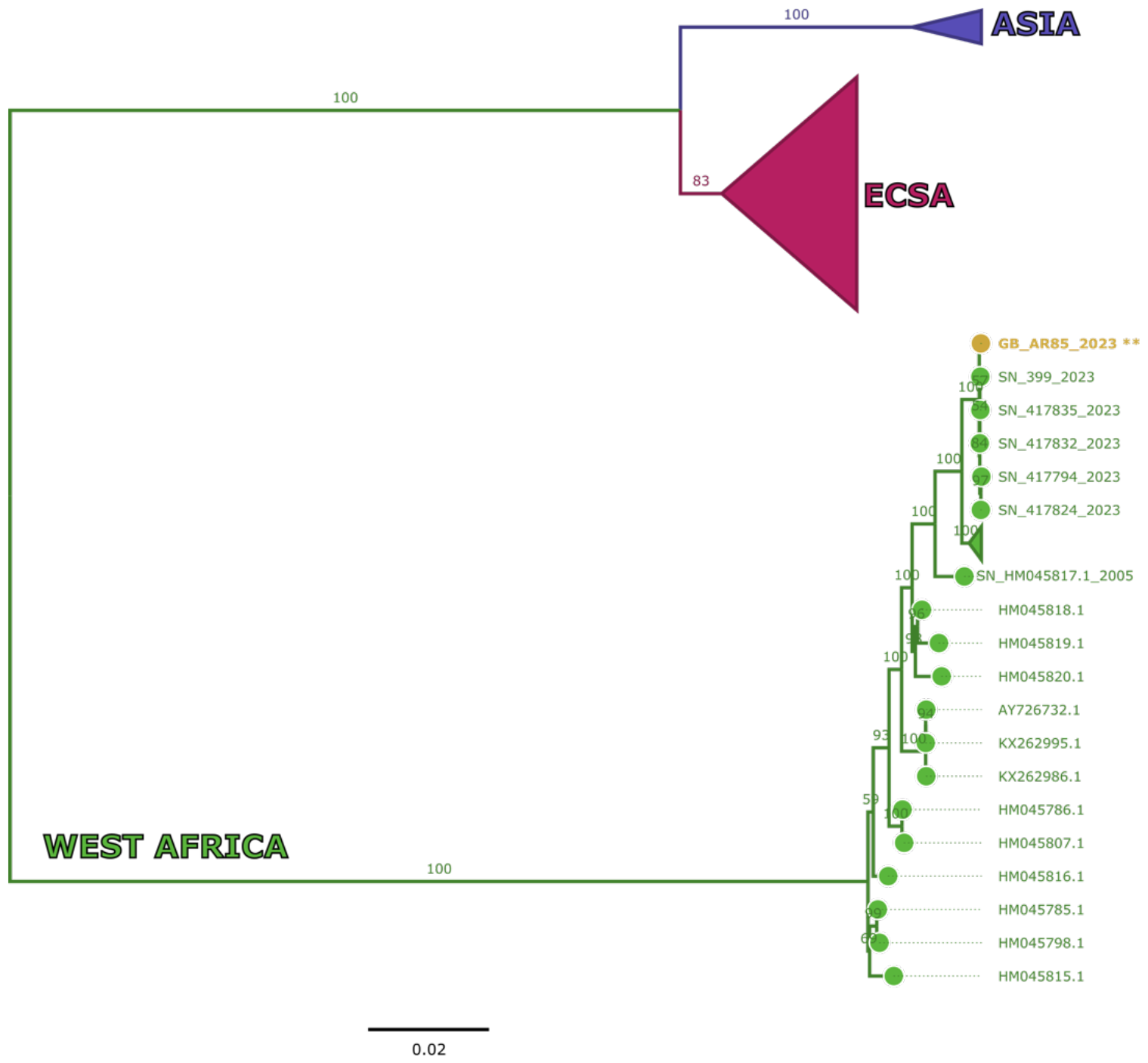
Maximum Likelihood (ML) tree depicted the relationship of sequenced Gambian CHIKV cases with global sequences. Viruses belonging to West African genotype are colored in green; Gambian sequence is colored in yellow. Patient sequences identifying number (AR85) is anonymized and unknown outside lab staff members.

To our knowledge, this is the first report of an autochthonous CHIKV infection ever identified in The Gambia. Genomic analysis highlight that the CHIKV emergence in The Gambia is linked to the ongoing outbreak in Senegal, indicating the viral strain’s spread across both countries’ borders. Previous evidence of circulation of arboviruses between the two countries has already been reported in the past. Indeed, in 2018, a serologically confirmed fatal case of Rift Valley fever had already been reported after a round trip between Gambia and Senegal (7). In the same year, the 2020-2021 Yellow fever outbreak in eastern Senegal was linked to a strain detected in 2018 from a Dutch traveler who fell ill after staying in Gambia and Senegal (8).

Urban development and increased trade lead to economic and societal gains, but also trigger potential changes in arboviruses epidemiology necessitating a cross-country collaboration and mitigation strategies. On the other hand, biological, socioeconomic, and ecological changes increase the risk of viral respiratory infections emergence and transmission worldwide (11).

The dual challenge of climate change and pandemics necessitates a comprehensive surveillance approach for (re-)emerging infectious diseases. The syndromic sentinel surveillance system showed its efficiency as a early warning system and outbreak response in Senegal (12,13).

## Ethical considerations

The Gambian National Ethical Committee of the Ministry of Health and Social Welfare approved the surveillance protocol which led to the obtention of human sera as less minimal risk research, and written consents were not required. Throughout the study, the database was shared with the Service Ministry of Health and Social Welfare of the Gambia for appropriate public health action.

## Data Availability

All data produced in the present study are available upon reasonable request to the authors

## Acknowledgment

We would like to convey special thanks to all the worker of the Epidemiology and Disease Control Department, the Ministry of Health and Social Welfare of The Gambia, the Virology department of the Institut Pasteur de Dakar and the National Public Health Laboratories of The Gambia

---

**Figure S1.**
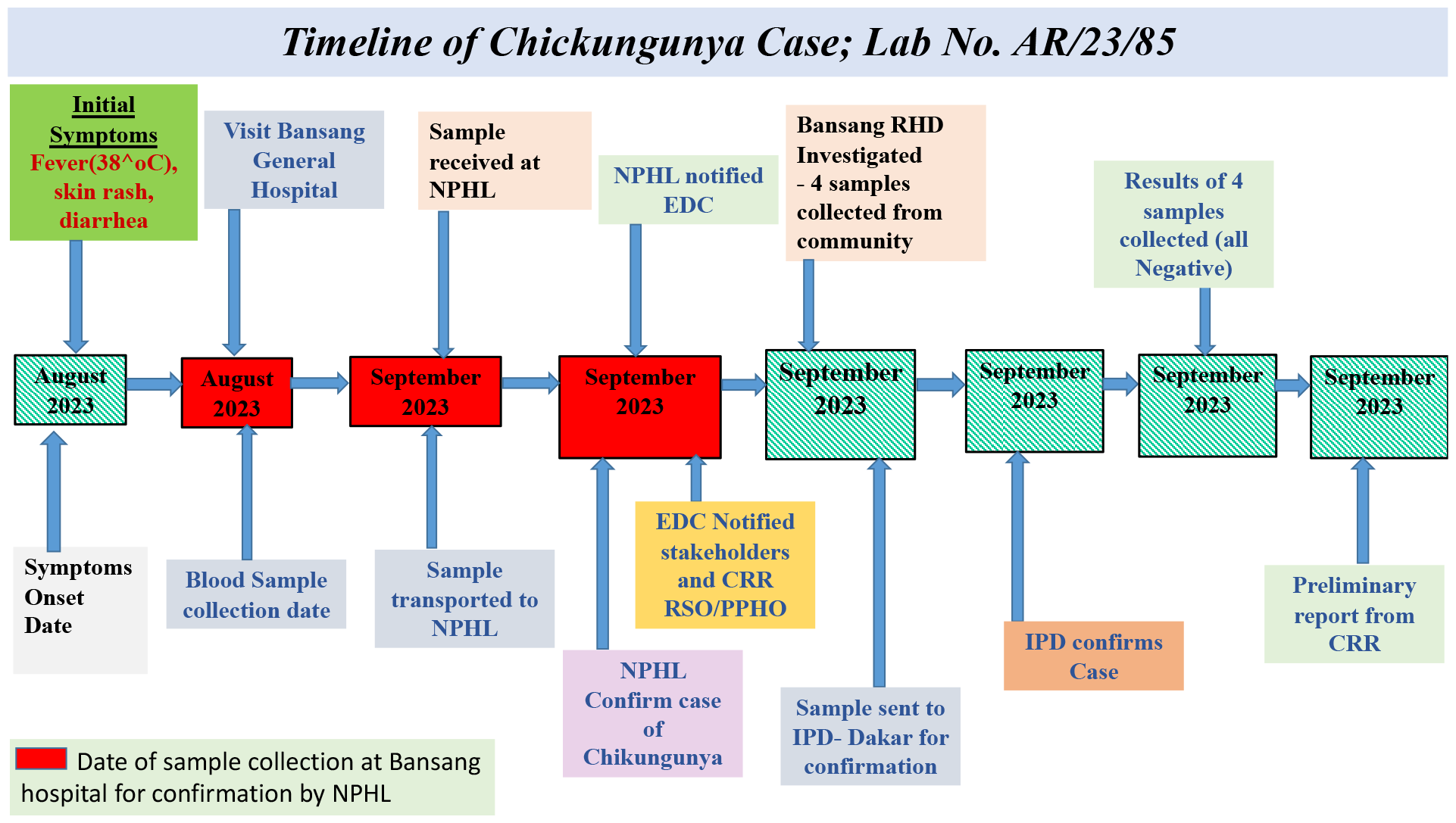
Timeline of Key events related to detected CHIKV case

## References

1. Morens DM, Fauci AS. Emerging infectious diseases: threats to human health and global stability. PLoS Pathog. 2013;9(7):e1003467.

2. Dia N, Diene Sarr F, Thiam D, Faye Sarr T, Espié E, OmarBa I, et al. Influenza-Like Illnesses in Senegal: Not Only Focus on Influenza Viruses. PLoS ONE [Internet]. 27 mars 2014 [cité 24 oct 2019];9(3). Disponible sur: https://www.ncbi.nlm.nih.gov/pmc/articles/PMC3968133/

3. Bob NS, Bâ H, Fall G, Ishagh E, Diallo MY, Sow A, et al. Detection of the Northeastern African Rift Valley Fever Virus Lineage During the 2015 Outbreak in Mauritania. Open Forum Infect Dis [Internet]. 2017 [cité 30 août 2019];4(2). Disponible sur: https://academic.oup.com/ofid/article-lookup/doi/10.1093/ofid/ofx087

4. Wu L, Mwesigwa J, Affara M, Bah M, Correa S, Hall T, et al. Antibody responses to a suite of novel serological markers for malaria surveillance demonstrate strong correlation with clinical and parasitological infection across seasons and transmission settings in The Gambia. BMC Med. 25 sept 2020;18(1):304.

5. Ikumapayi UN, Hill PC, Hossain I, Olatunji Y, Ndiaye M, Badji H, et al. Childhood meningitis in rural Gambia: 10 years of population-based surveillance. Mossong J, éditeur. PLOS ONE. 10 août 2022;17(8):e0265299.

6. Idrissa Dieng, Bacary Djilocalisse Sadio, Alioune Gaye, Samba Niang Sagne, Ndione MHD, Kane M, et al. Genomic characterization of a reemerging Chikungunya outbreak in Kedougou, Southeastern Senegal, 2023. 2023 [cité 4 mars 2024]; Disponible sur: 10.13140/RG.2.2.32038.50249

7. Rift Valley fever – Gambia [Internet]. [cité 4 mars 2024]. Disponible sur: https://www.who.int/emergencies/disease-outbreak-news/item/26-february-2018-rift-valley-fever-gambia-en

8. Diagne MM, Ndione MHD, Gaye A, Barry MA, Diallo D, Diallo A, et al. Yellow Fever Outbreak in Eastern Senegal, 2020–2021. Viruses. 28 juill 2021;13(8):1475.

9. Longbottom J, Walekhwa AW, Mwingira V, Kijanga O, Mramba F, Lord JS. Aedes albopictus invasion across Africa: the time is now for cross-country collaboration and control. Lancet Glob Health. avr 2023;11(4):e623–8.

10. Wilder-Smith A, Lindsay SW, Scott TW, Ooi EE, Gubler DJ, Das P. The Lancet Commission on dengue and other Aedes-transmitted viral diseases. The Lancet. juin 2020;395(10241):1890–1.

11. 1. He Y, Liu WJ, Jia N, Richardson S, Huang C. Viral respiratory infections in a rapidly changing climate: the need to prepare for the next pandemic. eBioMedicine. juill 2023;93:104593.

12. Dieng I, Barry M, Diagne M, Diop B, Ndiaye M, Faye M, et al. Detection of Crimean Congo haemorrhagic fever virus in North-eastern Senegal, Bokidiawé 2019. Emerg Microbes Infect. 2020;

13. Ndione MHD, Ndiaye EH, Faye M, Diagne MM, Diallo D, Diallo A, et al. Re-Introduction of West Nile Virus Lineage 1 in Senegal from Europe and Subsequent Circulation in Human and Mosquito Populations between 2012 and 2021. Viruses. 6 déc 2022;14(12):2720.

